# Maternal diabetes and overweight as risk factors for congenital heart defects in offspring - A nationwide register study from Finland

**DOI:** 10.1101/2023.02.14.23285825

**Authors:** R Turunen, A Pulakka, J Metsälä, T Vahlberg, T Ojala, M Gissler, E Kajantie, E Helle

## Abstract

**Importance:** Congenital heart defects (CHDs) affect 1–2% of newborns and are associated with significant mortality and morbidity. Understanding risk factors underlying CHDs is essential for prevention.

**Objective:** To determine the association between maternal diabetes and overweight/obesity and CHDs among offspring.

**Design:** Nationwide population-based register study.

**Setting:** Finland

**Participants:** All children born between 2006–2016 (N=620 751), and their mothers.

**Exposures:** Maternal pre-pregnancy body mass index (BMI) categorized as underweight (<18.5 kg/m^2^), normal (18.5–24.9 kg/m^2^), overweight (25.0–29.9 kg/m^2^), and obese (≥30 kg/m^2^). Maternal diabetes classified as no diabetes, type 1 (T1DM), type 2/other (T2DM), and gestational diabetes (GDM).

**Main Outcomes and Measures:** Odds ratio (OR) of isolated CHD in the child. In addition, nine anatomical CHD subgroups were studied.

**Results:** Of the 620 751 children born in Finland during the study period, 10 254 (1.65%) had an isolated CHD. T1DM was associated with an increased risk of having a child with any CHD (OR 3.71 (95% CI 3.16–4.35)), whereas maternal overweight (OR 0.98 (95% CI 0.98–1.04)) and obesity (OR 1.00 (95% CI 0.93–1.07)) were not. When analyzing anatomical subgroups, T1DM was associated with an increased risk in six subgroups. Maternal overweight was associated with complex defects (OR 2.24 (95% CI 1.01–4.94)), left ventricular outflow tract obstruction (OR 1.26, (95% CI 1.07–1.49), maternal obesity with complex defects (OR 3.22 (95% CI 1.31–7.92)), and right ventricular outflow tract obstruction (OR 1.26, (95% CI 1.01–1.55)). At the population level, maternal diabetes was responsible for 3.0% and maternal overweight and obesity for 0.7% of offspring’s CHD.

**Conclusions and Relevance:** This study indicated a less profound association between maternal overweight and obesity and CHD in the offspring than previously reported. The different risk profiles of T1DM and overweight/obesity may suggest distinct underlying teratogenic mechanisms.

## Introduction

Congenital heart defects (CHDs) are the most common congenital malformations in children, traditionally thought to affect roughly one in a hundred newborns ^1^. Together with prematurity and birth asphyxia and trauma, they are among the leading cause of deaths in the first year of life in high-resource settings ^2^. According to the US Center for Disease Control and Prevention, CHDs accounted for the largest percentage of birth defect associated hospitalizations (14.0%), and the highest total cost, approximately $6.1 billion (26.6% of total birth defect–associated hospitalization costs) in 2013 ^3^. Even though most children with CHDs survive to adulthood, CHDs are associated with significant mortality, morbidity, and reduced quality of life ^4,5^. It has been estimated that nearly 12 million people were living with CHDs globally in 2017 ^2^.

While there clearly is a hereditary component associated with CHD, potentially modifiable maternal factors, such as maternal overweight, obesity, and pregestational and gestational diabetes mellitus (PGDM and GDM) have been associated with increased risk for CHD in offspring ^6–10^. The role of maternal type 1 diabetes mellitus (T1DM), as a significant risk factor has been well documented, but the significance of maternal obesity and overweight is less clear. A better understanding of the contribution of maternal diabetes and obesity to an offspring’s CHD risk would not only aid prevention, but also provides cues to direct research in unraveling the underlying molecular-level mechanisms. We studied the association of maternal PGDM and GDM as well as overweight and obesity as risk factors for isolated CHD and selected CHD subgroups in a nationwide register study from Finland.

## Methods

### Study design

We conducted a nationwide register study in Finland including all children born (liveborn and stillborn) in 2006–2016 and their mothers (eFigure1). Exclusion criteria included missing information on gestational age, unclear CHD diagnosis, and having diagnosis for syndromes, chromosomal aberrations, and extracardiac malformations (major anomalies according to EUROCAT)^11^.

### Data sources

Data was collected from the following national registers: Medical Birth Register (MBR), Register of Congenital Malformations (RCM), and Care Register for Health Care (CRHC) maintained by the Finnish Institute for Health and Welfare (THL); and the Register of Special Reimbursements for Prescription Medicines maintained by the Social Insurance Institution of Finland (SII)^12,13^. Every Finnish citizen and permanent resident has a personal identification number, which enables linkage of information between different registers. The information on maternal and paternal education was received from Statistics Finland. A detailed description of these registers is provided in eMethods.

### Outcomes

The main outcome was isolated CHD in the child obtained from the RCM. Isolated CHD was defined as having a diagnosis for one or more CHD and not having diagnoses for chromosomal aberrations, syndromes, or any other major extracardiac anomalies (eTable 1). Isolated CHDs were divided into nine groups according to their anatomical origin: complex, transposition of great arteries (TGA), left ventricular outflow tract obstruction (LVOTO), right ventricular outflow tract obstruction (RVOTO), pulmonary venous anomalies, anomalies of thoracic arteries and veins, atrial septal defects (ASD), ventricular septal defects (VSD), and other septal defects (eTable 2).

**Table 1.**
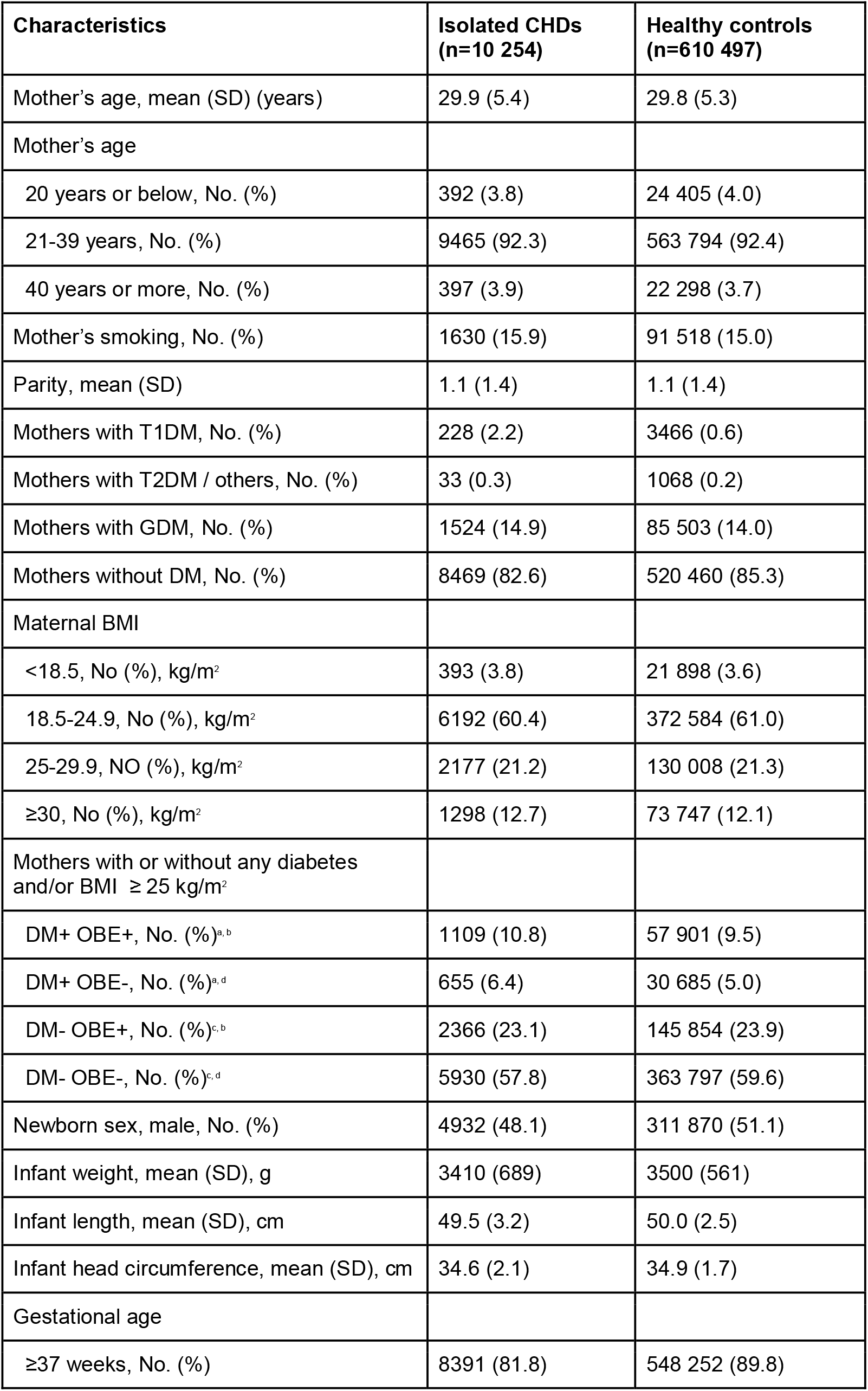
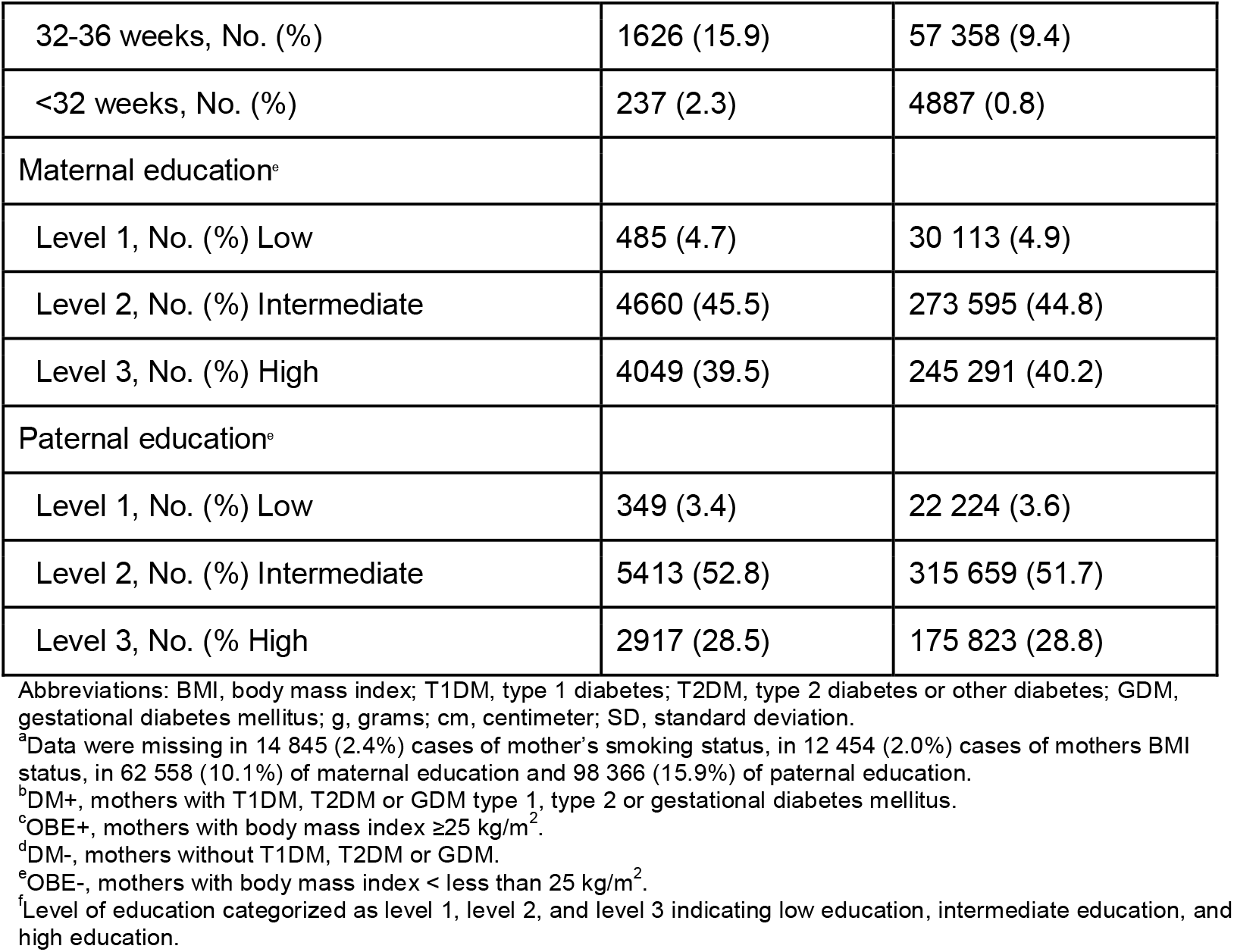
Baseline characteristics of the individuals in the study (N=620 751)

**Table 2.**
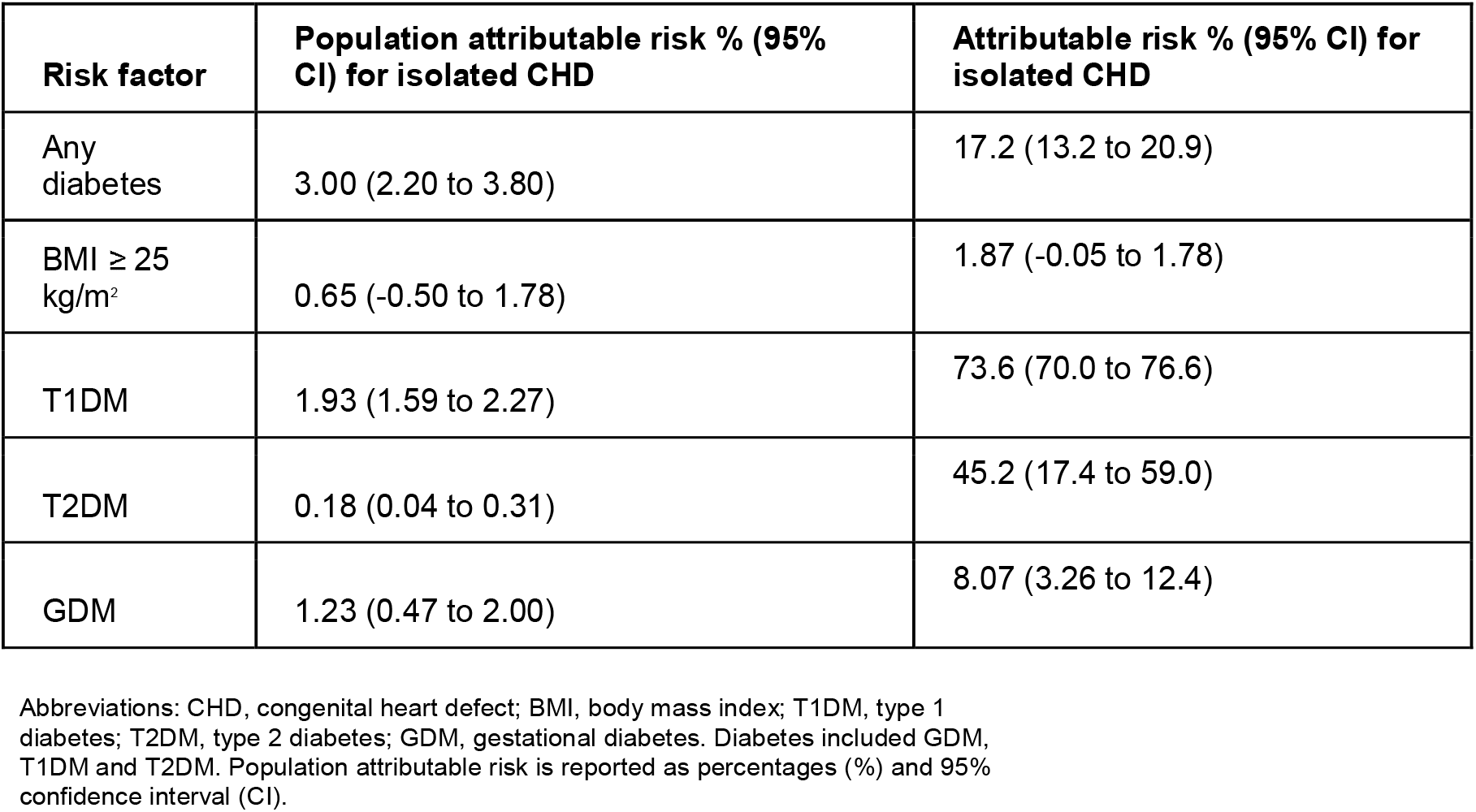
Population attributable risk for isolated CHD when exposed to maternal diabetes and overweight or obesity

| **Risk factor** | **Population attributable risk % (95% CI) for isolated CHD** | **Attributable risk % (95% CI) for isolated CHD** |
| --- | --- | --- |
| Any diabetes | 3.00 (2.20 to 3.80) | 17.2 (13.2 to 20.9) |
| BMI ≥ 25 kg/m<sup>2</sup> | 0.65 (-0.50 to 1.78) | 1.87 (-0.05 to 1.78) |
| T1DM | 1.93 (1.59 to 2.27) | 73.6 (70.0 to 76.6) |
| T2DM | 0.18 (0.04 to 0.31) | 45.2 (17.4 to 59.0) |
| GDM | 1.23 (0.47 to 2.00) | 8.07 (3.26 to 12.4) |

Abbreviations: CHD, congenital heart defect; BMI, body mass index; T1DM, type 1 diabetes; T2DM, type 2 diabetes; GDM, gestational diabetes. Diabetes included GDM, T1DM and T2DM. Population attributable risk is reported as percentages (%) and 95% confidence interval (CI).

### Exposures and covariates

Maternal body mass index (BMI) was calculated from the weight and height reported in the MBR. Height was self-reported. Pre-pregnancy weight is self-reported during the first antenatal clinic visit, when weight is also measured. BMI was categorized into underweight (<18.5 kg/m^2^), normal (18.5–24.9 kg/m^2^), overweight (25.0–29.9 kg/m^2^), and obese (≥30.0 kg/m^2^). Maternal diabetes was classified into no diabetes, T1DM, type 2 other (T2DM), and GDM according to the information in MBR, CRHC, and Register of Special Reimbursements for Prescription Medicines (eMethods).

The covariates considered as potential confounders included year of birth, maternal age, parity, maternal smoking during pregnancy (yes/no), and offspring sex (M/F) obtained from the MBR, and highest maternal and paternal level of education (eMethods) obtained from Statistics Finland.

### Statistical analysis

Categorical variables were reported as frequencies and percentages, and continuous variables as means and standard deviations. Logistic regression adjusted for child’s birth year was used for analyzing the association between maternal BMI classes and diabetes on isolated CHD and selected CHD subgroups in the offspring. Additionally, multivariable logistic regression analysis was performed for maternal DM and BMI, adjusted for the above mentioned covariates. Results of logistic regression are presented as odds ratios (ORs) with 95% confidence intervals (CI) as measure of associations. Those with missing data were excluded from the analysis. The comparison of isolated CHD frequencies between those with missing and non-missing data was made with chi-square test.

Population attributable risk (PAR) was calculated using an indirect method to analyze the risk for isolated CHD in subjects of the total population that are attributable to risk factors, which were maternal diabetes and obesity. Two-sided p-values < .05 were considered statistically significant. Data were analyzed using SAS Enterprise Guide version 8.3 (SAS Institute, Cary, North Carolina).

### Ethical considerations

The study was approved by the Research Ethics board of the Finnish Institute for Health and Welfare (THL/1960/6.02.01/2018,§810) and relevant register authorities. No informed consent from the registered persons is required for the use of pseudonymised register data for research purposes in Finland.

## Results

The study population consisted of 620 751 children, of whom 10 254 (1.65%) had an isolated CHD (eTable 4). The baseline characteristics of the individuals in the study and maternal pregnancy data are presented in Table 1.

The prevalence of GDM increased from 10.3% to 19.2% during the study period, whereas T2DM increased from 0.1% to 0.3%, and the prevalence of T1DM was stable at approximately 0.7% (eFigure 2). Maternal overweight increased from 20.3% to 22.2% and maternal obesity from 10.7% to 13.3% (eFigure 3). GDM was highly correlated with BMI in the CHD mothers, as 8.2% of the normal weight mothers, 21.1% of the overweight mothers, and 41.1% of the obese mothers had a GDM diagnosis (eTable 5).

**Figure 1.**
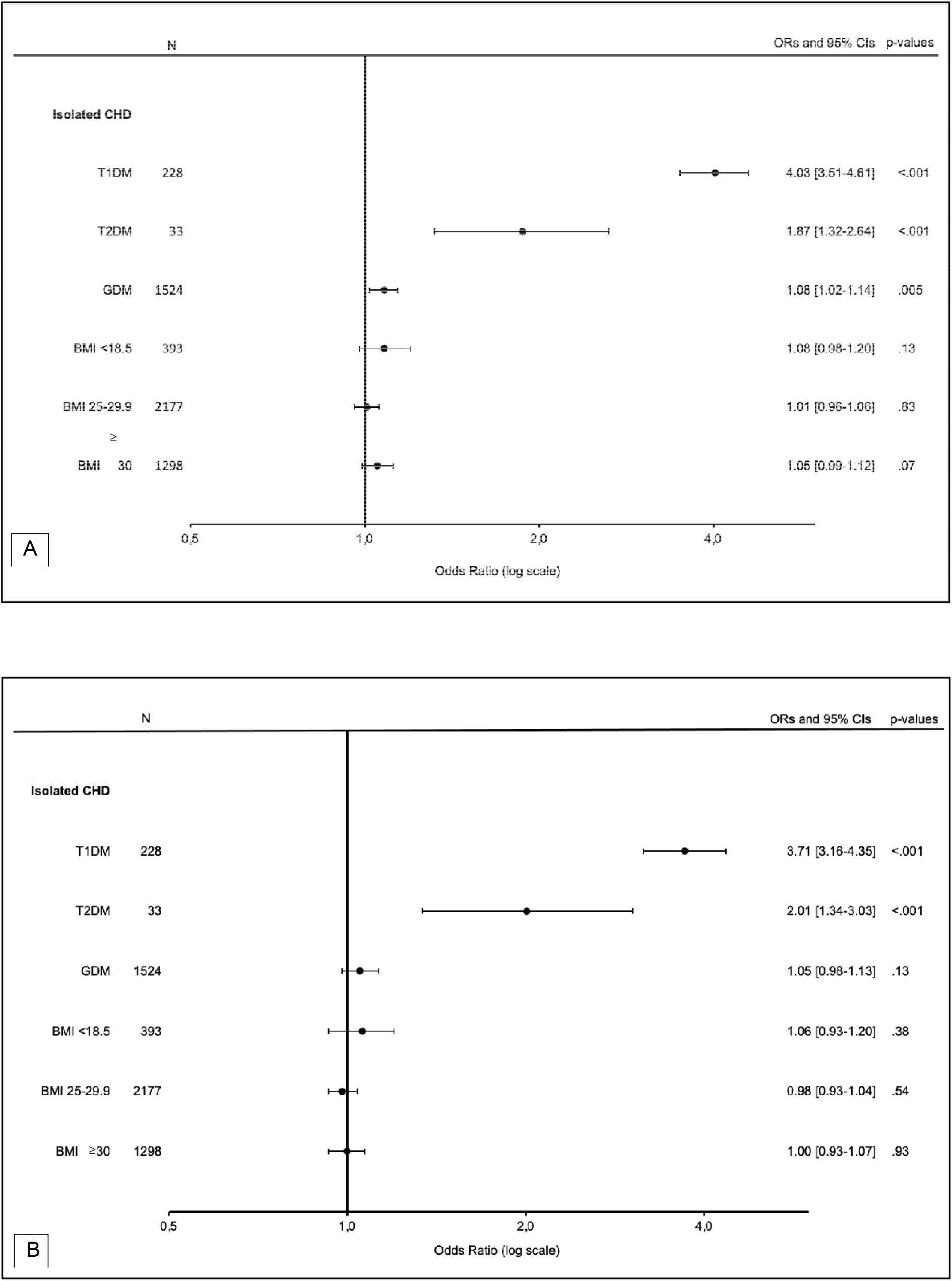
The association between maternal BMI and diabetes and isolated CHD^a^ in offspring Abbreviations: CHD, congenital heart defect; BMI, body mass index; GDM, gestational diabetes mellitus; T1DM, type I diabetes; T2DM, type II diabetes; OR, odds ratio; CI, confidence interval. ^a^The association was analyzed using child’s birth year adjusted (A) and multivariable (B) logistic regression analysis. Normal BMI (18.5-24.9) and no DM were used as a reference group. The statistical significance was reached with p-value <.05.

**Figure 2.**
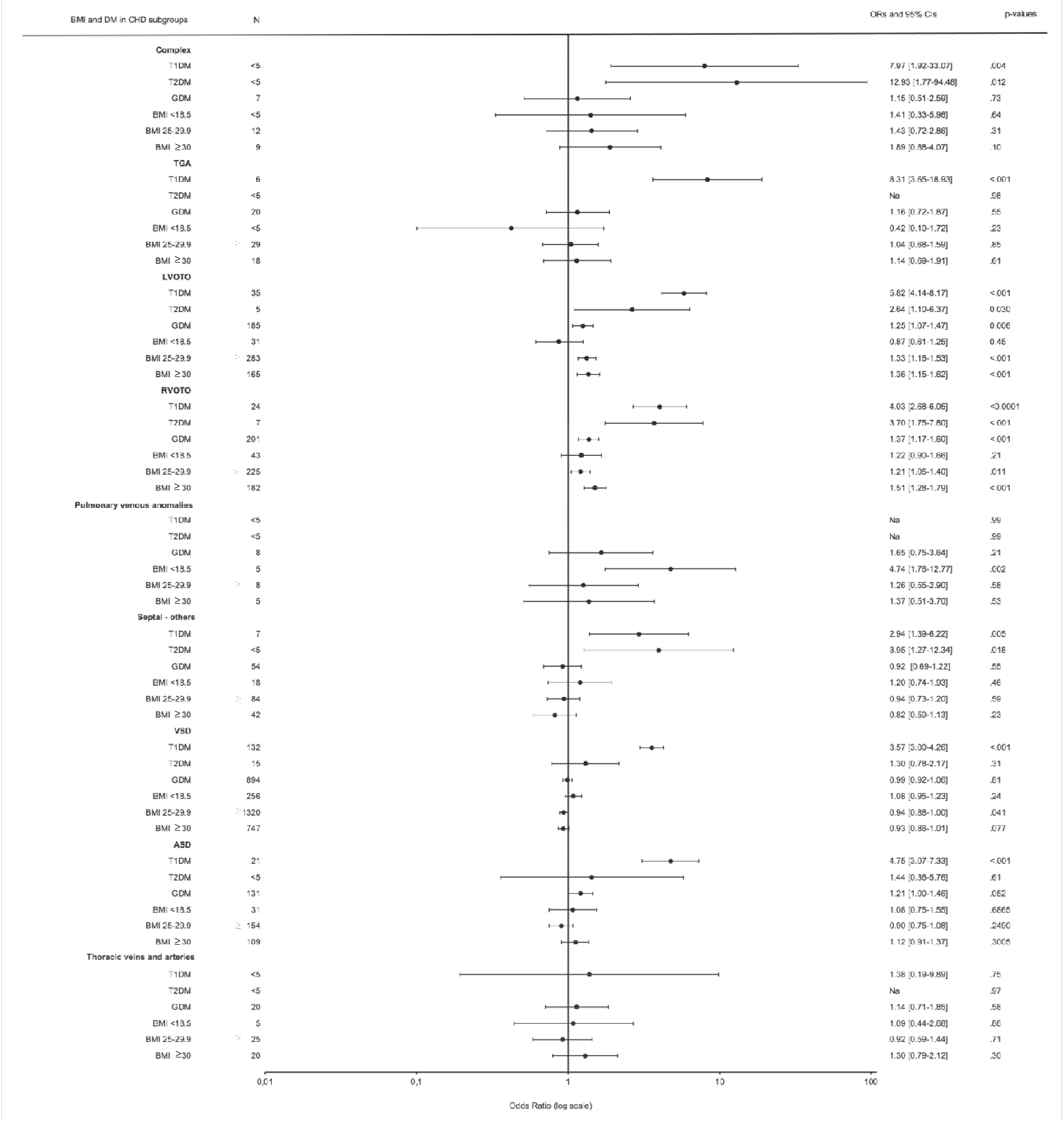
The association between maternal BMI and diabetes and CHD subgroups in offspring^a^ Abbreviations: CHD, congenital heart defects; BMI, body mass index; GDM, gestational diabetes mellitus; T1DM, type 1 diabetes; T2DM, type 2 diabetes or other diabetes; Na, not able to analyse; OR, odds ratio; CI, confidence interval; TGA, transposition of great arteries; LVOTO, left ventricle outflow tract obstruction; RVOTO, right ventricle outflow tract obstruction; VSD, ventricular septal defect; ASD, atrium septal defect. ^a^The analysis was adjusted to birth year of the child. Normal BMI (18.5-24.9) and no DM were used as a reference group. The statistical significance was reached with p-value <.05.

**Figure 3.**
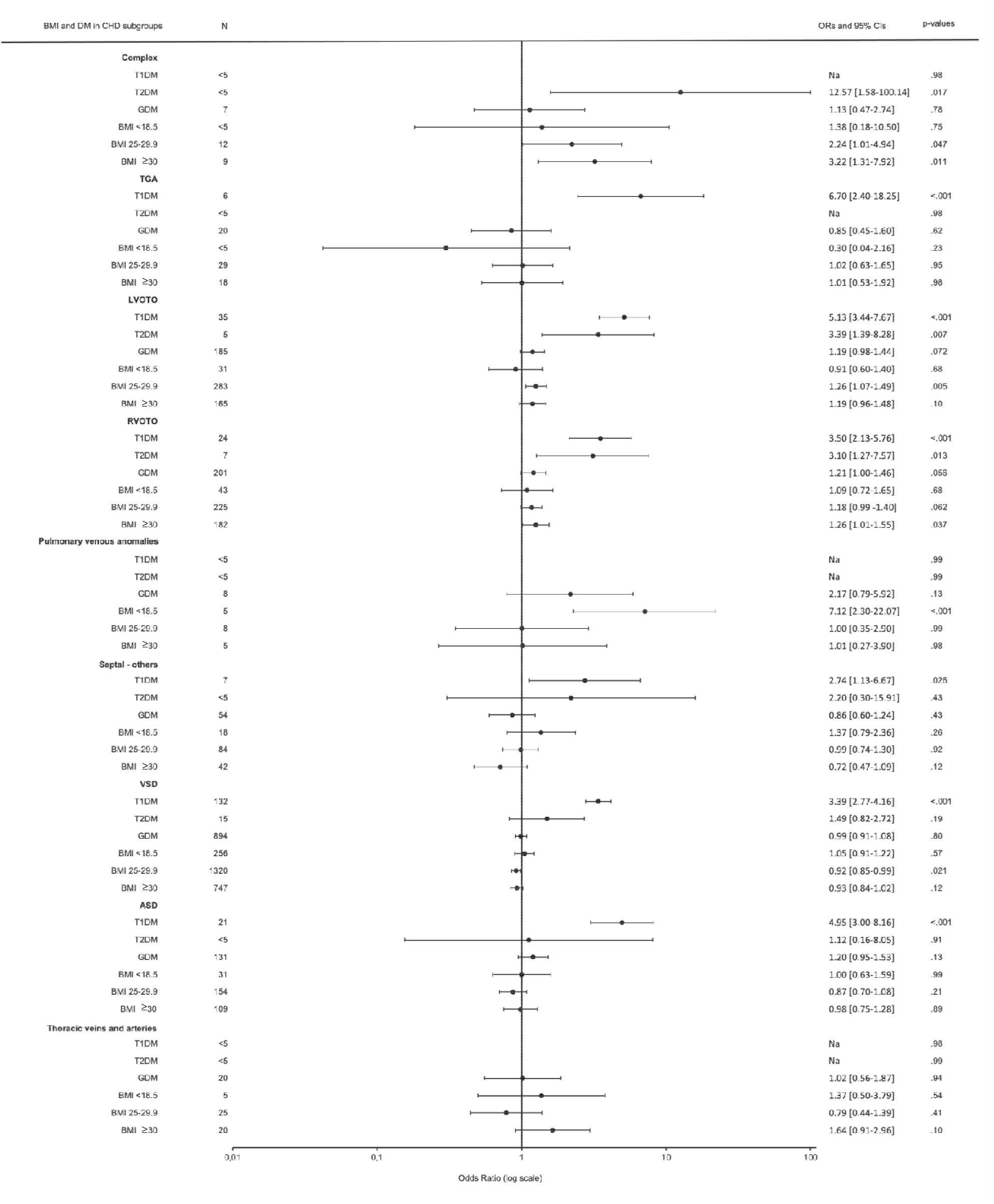
The association between maternal BMI and diabetes and CHD subgroups using multivariable logistic regression analysis^a^ Abbreviations: CHD, congenital heart defect; BMI, body mass index; GDM, gestational diabetes mellitus; DMI, type I diabetes; DMII, type II diabetes; Na, not able to analyse; OR, odds ratio; CI, confidence interval; TGA, transposition of great arteries; LVOTO, left ventricle outflow tract obstruction; RVOTO, right ventricle outflow tract obstruction; VSD, ventricular septal defect; ASD, atrium septal defect. ^a^The analysis was adjusted to maternal smoking, child sex, maternal age, birth year of the child, first parity, maternal and parental education. Normal BMI (18.5-24.9) and no DM were used as a reference group. The statistical significance was reached with p-value <.05.

Logistic regression adjusted for child’s birth year indicated increased risk between maternal T1DM (OR 4.03 (95% CI 3.51–4.61)), T2DM (OR 1.87 (95% CI 1.32–2.64)) or GDM ((OR 1.08 (95% CI 1.02–1.14)) and isolated CHD in the offspring, while the associations between overweight ((OR 1.01 (95% CI 0.96–1.06)) and obesity ((OR 1.05 (95% CI 0.99–1.12)) and isolated CHD in the offspring were not statistically significant (Figure 1a). In a multivariable logistic regression analysis maternal T1DM (OR 3.71 (95% CI 3.16–4.35)) and T2DM (OR 2.01 (95% CI 1.34–3.03)) remained significantly associated with an increased risk (Figure 1b). We found no association between maternal obesity (OR 1.00 (95% CI 0.93–1.07)) and overweight (OR 0.98 (95% CI 0.98–1.04)) and isolated CHD in the offspring in the multivariable logistic regression analysis, when the CHD were analyzed as a single group.

### CHD subgroup analyses

The logistic regression analysis with offspring’s CHD subgroups as outcomes, adjusted for birth year, is presented in Figure 2. After adjusting the models for the other covariates (Figure 3), T1DM was clearly the strongest risk factor for TGA (OR 6.70 (95% CI 2.40–18.25), LVOTO (OR 5.13 (95% CI 3.44–7.67)), RVOTO (OR 3.50 (95% CI 2.13–5.76)) and isolated ASD (OR 4.95 (95% CI 3.00–8.16) and VSD (OR 3.39 (95% CI 2.77–4.16)) as well as other septal defects (OR 2.74 (95% CI 1.13–6.67)). Maternal overweight was associated with an increased risk for complex defects (OR 2.24 (95% CI 1.01–4.94)) and LVOTO (OR 1.26 (95% CI 1.07–1.49)), compared with normal weight. Maternal obesity was associated with an increased risk for complex defects (OR 3.22 (95% CI 1.31–7.92)) and RVOTO (OR 1.26 (95% CI 1.01–1.55)). Finally, maternal overweight was associated with a lower risk for VSD (OR 0.92 (95% CI 0.85–0.99)) in the offspring, and maternal underweight (BMI <18.5 kg/m^2^) was associated with an increased risk for pulmonary venous anomalies (OR 7.12 (95% CI 2.30–22.07)).

### Population attributable risk

We determined the population attributable risk with overweight/obesity and diabetes on having CHD in the offspring (Table 2). The attributable risk of diabetes was 17.20% (95% CI 13.20 to 20.90), i.e. 17% of the risk of having offspring with CHD in the maternal diabetes group is attributed to any diabetes. The population attributable risk of any diabetes was 3.00% (95% CI 2.20% to 3.80%), i.e. 3% of the risk of having offspring with CHD in the whole population is attributed to any diabetes. The attributable risk of overweight and obesity was 1.87% (95% CI −0.05% to 1.78%), and population attributable risk was 0.65% (95% CI −0.50% to 1.78%), but these associations did not reach statistical significance.

### Missing data

Maternal BMI status was missing in 2.01% of the individuals. Mother’s smoking status during pregnancy was missing in 14 845 (2.4%) cases, mother’s BMI in 12 454 (2.0%) cases, maternal education in 62 558 (10.1%) cases, and paternal education in 98 366 (15.9%) cases. There were no differences in the outcomes for those with missing and non-missing data (eTable 6).

## Discussion

We conducted a nationwide register study to quantify the effect of maternal diabetes and overweight/obesity in pregnancy on the risk for CHD in offspring. We show that maternal T1DM constitutes by the far the strongest risk, with 3.7-fold odds for any CHD. T2DM was associated with 2.0-fold odds, and GDM, overweight and obesity were associated with modest, approximately 1.1- to 1.2-fold odds. T1DM was associated with a significantly increased risk in most of the CHD subcategories, whereas maternal overweight and obesity were associated with increased risk for complex defects and slightly increased risk for LVOTO and RVOTO defects. In general, our study indicated weaker associations between maternal overweight and obesity and CHD in the offspring than previously reported, and we speculate that this is due to our comprehensive data on maternal diabetes, which likely accounts for a larger part of the risk in overweight and obese individuals than previously thought.

Finally, our results may suggest that maternal diabetes and overweight/obesity have distinct teratogenic mechanisms, as the associations were different for many of the CHD subgroups, in some cases even opposite.

The association of maternal T1DM with offspring risk for CHD is well known. In line with previous studies ^9,14–16^, our results demonstrated a three to four-fold increased risk for any CHD, and the risk was increased for all the CHD subgroups that had enough cases to determine it. GDM was associated with a slightly increased risk for any CHD and LVOTO defects in the logistic regression adjusted for child’s birth year, however these associations were not statistically significant in the multivariable analysis. As GDM is highly prevalent — affecting over one in three expectant mothers — any risk increase is important at the population level. This was demonstrated by a population attributable risk of 1.23% for GDM compared with 1.93% for T1DM, which is considerably less prevalent.

During the 2006–2016 study period, the screening policy of GDM was changed from risk factor based to comprehensive screening, introduced by a 2008 guideline. This resulted in an increased number of women screened and milder cases included as GDM with on average less severe perinatal and neonatal outcomes ^17,18^. In addition, the proportion of obese and older age (≥35 years) parturients increased during the study period ^19^, which may also underlie the increase in GDM prevalence. These changes are also likely to affect the association between diagnosed GDM and CHDs.

Maternal obesity and overweight were associated with a less profound risk for offspring’s CHD in our study than previously reported, when analyzed together with maternal diabetes. This is in line with a recent smaller study from China, showing that PGDM partially mediated the association between maternal obesity and CHD ^20^. In our study, maternal overweight and obesity were associated with an increased risk for complex defects and outflow tract obstruction defects, whereas two large previous studies have indicated association with a wider range of defects and with higher odds ratios ^6,8^. These studies, however, did not adjust for maternal diabetes comprehensively. The study by Persson et al. on 28 628 individuals with CHD and 2 050 491 controls excluded subjects with PGDM from the analysis, and lacked GDM as a covariate in their multivariate model ^8^. The study by Madsen et al., including 11 263 individuals with CHD and 140 470 controls, presented no information on T1DM^6^. Although they did adjust the analyses for GDM, the prevalence of GDM was 6.3% in cases and 4.3% in controls, and thus GDM was potentially underreported. In our study GDM was highly correlated with BMI in CHD mothers. Thus, it is likely that these studies underestimated the role of maternal diabetes in their analyses.

In several smaller studies showing no or some association with maternal BMI and offspring’s CHD the association with specific CHD subtypes has been inconsistent ^7,21–24^. Indeed, analyses of CHD subtypes are limited by low prevalence of individual malformations, even in large cohorts. A recent meta-analysis including 19 studies with 2,416,546 participants reported pooled relative risks of having infants with CHDs of 1.08 (95% CI 1.03–1.13) in overweight and 1.23 (95% CI 1.17–1.29) in obese mothers. Nonetheless, most included studies were case-control studies and few adjusted for confounding factors such as maternal age, smoking, education, or diabetes ^25^. Moreover, in many of the studies BMI was based on retrospective self reported data, which is subject to recall bias.

Interestingly, our data indicated a lower risk for VSD in offspring of overweight mothers while previous reports have shown no association ^8^ or increased risk ^7^. Maternal obesity has been shown to be associated with increased birth weight in the offspring ^26,27^, and increased left ventricle ^28^ and interventricular septum ^29^ thickness during infancy. Thus, one explanation could be that the increased septum thickness in these infants might contribute to the closure of small muscular defects before they are diagnosed. On the other hand, maternal T1DM is also known to be associated with ventricular hypertrophy in the offspring in the neonatal period ^30^, but unlike maternal overweight, it is also associated with an increased risk for offspring’s VSD as shown by us and others. This suggests that whereas T1DM is associated with abnormal cardiac septation, maternal overweight and obesity is not, pointing to different teratogenic mechanisms in these two conditions.

Higher plasma glucose values during early pregnancy have been associated with an increased risk for CHD in the offspring in non-diabetic mothers ^31^, and hyperglycemia is likely the major teratogenic factor in T1DM ^32,33^. The pathophysiological processes behind GDM and obesity-associated risk are less well known. It is not unreasonable to speculate that at least some mothers diagnosed with GDM later in pregnancy have glycemic dysregulation and pathologically high glucose values already in early pregnancy, however, additional mechanisms should be considered. Recent studies have demonstrated distinct early pregnancy metabolomic profiles, including exaggerated dyslipidemia and increased inflammatory markers in mothers who later develop GDM ^34–36^. Abnormal early pregnancy maternal lipid profiles have been associated with increased risk for CHD in the offspring ^37,38^. Both obesity and GDM are associated with increased oxidative stress and endothelial dysfunction, and endocardial dysfunction has been proposed as one pathophysiological origin of LVOTO defects ^39–41^. These findings suggest that obesity and GDM-mediated abnormal metabolomics related to inflammation, oxidative stress, and hyperlipidemia, could be associated with an additive risk in genetically predisposed individuals. The association between maternal overweight and obesity as a risk for LVOTO, RVOTO, and complex defects warrants further mechanistic research on these CHD subtypes to identify potentially modifiable pathophysiological processes.

### Strengths and limitations

This study is thus far the largest population-based study to address the combined effect of diabetes and obesity on offspring’s risk for CHD. Although we used a national 11-year cohort, the number of study subjects in the CHD subgroups remains limited, which was seen as large confidence intervals associated with many of the significant findings. Another important limitation was that the data on pregnancy terminations and miscarriages was not reliably available. Congenital anomalies are known to be common in miscarriages and annually around 350 pregnancy terminations are made due to major congenital anomalies in Finland. It is likely that some associations were missed due to not having this data.

The strengths of this study include using nationwide register data with an unselected population of all children born in 2006–2016 in Finland. The data on exposures and outcomes were prospectively collected and comprehensively and reliably defined. Maternal diabetes was defined based on three unrelated registers, and we assessed the effect of PGDM and GDM separately. In addition, we were able to assess the severity of obesity. For the analysis we included only individuals with isolated heart defects without syndromes and any other major malformations leading to exclusion of most individuals with definitive or probable larger structural genetic defects with likely different etiologies. Thus, despite the limitations, this study and its findings demonstrate how combined data from several nationwide registers is an extremely valuable source for studying the origins of rare diseases.

### Conclusion

This study emphasizes T1DM as a risk factor for CHD whereas, at least in a high-resource setting with universal antenatal care, GDM and maternal overweight and obesity are associated with a much smaller risk. However, with increasing prevalence of GDM and maternal overweight, the risk at the population level is substantial. It has been shown that standard treatment of maternal DM results in significant reduction of the risk of anatomical malformations in offspring ^42^. Thus, primary prevention of overweight and obesity and careful treatment of PGDM hold the opportunity to reduce the burden of disease. Finally, a better understanding of the underlying mechanisms of maternal overweight and obesity in offspring’s increased risk for LVOTO, RVOTO, and complex defects could further improve the prevention of these CHD subtypes.

## Data Availability

According to Finnish legislation researchers working outside the statutory register authority Finnish Institute for Health and Welfare can access register data for research purposes by applying for permission from the Finnish Social and Health Data Permit Authority Findata (findata.fi/en)

## Conflict of Interest Disclosures

None reported

This study was supported by: Academy of Finland, Finnish Medical Foundation, Finnish Foundation for Pediatric Research, Finnish Foundation for Cardiovascular Research, European Commission (733280 RECAP Research on Children and Adults Born Preterm), Sigrid Juselius Foundation, Signe and Ane Gyllenberg Foundation, Yrjö Jahnsson Foundation, Novo Nordisk Foundation, Finnish Diabetes Research Foundation. This study was also supported by European Commission through Horizon 2020 (874739) and Horizon Europe (101057739).

## Role of the Funder/Sponsor

The funders had no role in the design and conduct of the study; collection, management, analysis, and interpretation of the data; preparation, review, or approval of the manuscript; and decision to submit the manuscript for publication.

## Notes

### Competing Interest Statement

The authors have declared no competing interest.

### Funding Statement

This study was supported by: Academy of Finland (331405) and (315690 NORDCAP), Finnish Medical Foundation, Finnish Foundation for Pediatric Research, Finnish Foundation for Cardiovascular Research, European Commission (733280 RECAP Research on Children and Adults Born Preterm), Sigrid Juselius Foundation, Signe and Ane Gyllenberg Foundation, Yrjo Jahnsson Foundation, Novo Nordisk Foundation, Finnish Diabetes Research Foundation. This study was also supported by European Commission through Horizon 2020 (874739) and Horizon Europe (101057739).

### Author Declarations

Ethics committee of the Finnish Institute for Health and Welfare gave ethical approval for this work

## References

1. Liu Y, Chen S, Zühlke L, et al. Global birth prevalence of congenital heart defects 1970–2017: updated systematic review and meta-analysis of 260 studies. Int J Epidemiol. 2019;48(2):455–463.

2. Zimmerman MS, Smith AGC, Sable CA, et al. Global, regional, and national burden of congenital heart disease, 1990–2017: a systematic analysis for the Global Burden of Disease Study 2017. The Lancet Child & Adolescent Health. 2020;4(3):185–200.

3. Hospital Costs for Birth Defects Reach Tens of Billions. JAMA. 2017;317(8):799.

4. Saha P, Potiny P, Rigdon J, et al. Substantial Cardiovascular Morbidity in Adults With Lower-Complexity Congenital Heart Disease. Circulation. 2019;139(16):1889–1899.

5. Calderon J, Stopp C, Wypij D, et al. Early-Term Birth in Single-Ventricle Congenital Heart Disease After the Fontan Procedure: Neurodevelopmental and Psychiatric Outcomes. J Pediatr. 2016;179:96–103.

6. Madsen NL, Schwartz SM, Lewin MB, Mueller BA. Prepregnancy Body Mass Index and Congenital Heart Defects among Offspring: A Population-based Study. Congenital Heart Disease. 2013;8(2):131–141. doi:10.1111/j.1747-0803.2012.00714.x

7. Brite J, Laughon SK, Troendle J, Mills J. Maternal overweight and obesity and risk of congenital heart defects in offspring. Int J Obes. 2014;38(6):878–882.

8. Persson M, Razaz N, Edstedt Bonamy AK, Villamor E, Cnattingius S. Maternal Overweight and Obesity and Risk of Congenital Heart Defects. J Am Coll Cardiol. 2019;73(1):44–53.

9. Øyen N, Diaz LJ, Leirgul E, et al. Prepregnancy Diabetes and Offspring Risk of Congenital Heart Disease: A Nationwide Cohort Study. Circulation. 2016;133(23):2243–2253.

10. Helle E, Priest JR. Maternal Obesity and Diabetes Mellitus as Risk Factors for Congenital Heart Disease in the Offspring. J Am Heart Assoc. 2020;9(8):e011541.

11. Greenlees R, Neville A, Addor MC, et al. Paper 6: EUROCAT member registries: Organization and activities. Birth Defects Research Part A: Clinical and Molecular Teratology. 2011;91(S1):S51–S100. doi:10.1002/bdra.20775

12. Laugesen K, Ludvigsson JF, Schmidt M, et al. Nordic Health Registry-Based Research: A Review of Health Care Systems and Key Registries. Clin Epidemiol. 2021;13:533–554.

13. Gissler M, Teperi J, Hemminki E, Meriläinen J. Data quality after restructuring a national medical registry. Scand J Soc Med. 1995;23(1):75–80.

14. Jenkins KJ, Correa A, Feinstein JA, et al. Noninherited Risk Factors and Congenital Cardiovascular Defects: Current Knowledge. Circulation. 2007;115(23):2995–3014. doi:10.1161/circulationaha.106.183216

15. Liu S, Joseph KS, Lisonkova S, et al. Association between maternal chronic conditions and congenital heart defects: a population-based cohort study. Circulation. 2013;128(6):583–589.

16. Hoang TT, Marengo LK, Mitchell LE, Canfield MA, Agopian AJ. Original Findings and Updated Meta-Analysis for the Association Between Maternal Diabetes and Risk for Congenital Heart Disease Phenotypes. American Journal of Epidemiology. 2017;186(1):118–128. doi:10.1093/aje/kwx033

17. Koivunen S, Torkki A, Bloigu A, et al. Towards national comprehensive gestational diabetes screening - consequences for neonatal outcome and care. Acta Obstet Gynecol Scand. 2017;96(1):106–113.

18. Koivunen S, Kajantie E, Torkki A, et al. The changing face of gestational diabetes: the effect of the shift from risk factor-based to comprehensive screening. Eur J Endocrinol. 2015;173(5):623–632.

19. Perinatal statistics – parturients, deliveries and newborns 2021. https://thl.fi/en/web/thlfi-en/statistics-and-data/statistics-by-topic/sexual-and-reproductive-health/parturients-deliveries-and-births/perinatal-statistics-parturients-delivers-and-newborns

20. Wu XX, Ge RX, Huang L, et al. Pregestational diabetes mediates the association between maternal obesity and the risk of congenital heart defects. J Diabetes Investig. 2022;13(2):367–374.

21. Cedergren MI, Källén BAJ. Maternal obesity and infant heart defects. Obes Res. 2003;11(9):1065–1071.

22. Block SR, Watkins SM, Salemi JL, et al. Maternal pre-pregnancy body mass index and risk of selected birth defects: evidence of a dose--response relationship. Paediatr Perinat Epidemiol. 2013;27(6):521–531.

23. Simeone RM, Tinker SC, Gilboa SM, et al. Proportion of selected congenital heart defects attributable to recognized risk factors. Ann Epidemiol. 2016;26(12):838–845.

24. Rutkowski RE, Tanner JP, Anjohrin SB, Kirby RS, Salemi JL. Proportion of critical congenital heart defects attributable to unhealthy prepregnancy body mass index among women with live births in Florida, 2005-2016. Birth Defects Res. 2021;113(18):1285–1298.

25. Liu X, Ding G, Yang W, et al. Maternal Body Mass Index and Risk of Congenital Heart Defects in Infants: A Dose-Response Meta-Analysis. Biomed Res Int. 2019;2019:1315796.

26. Ijäs H, Koivunen S, Raudaskoski T, Kajantie E, Gissler M, Vääräsmäki M. Independent and concomitant associations of gestational diabetes and maternal obesity to perinatal outcome: A register-based study. PLoS One. 2019;14(8):e0221549.

27. Alfadhli EM. Maternal obesity influences Birth Weight more than gestational Diabetes author. BMC Pregnancy Childbirth. 2021;21(1):111.

28. Guzzardi MA, Liistro T, Gargani L, et al. Maternal Obesity and Cardiac Development in the Offspring: Study in Human Neonates and Minipigs. JACC Cardiovasc Imaging. 2018;11(12):1750–1755.

29. Harink T den, den Harink T, Roelofs MJM, et al. Maternal obesity in pregnancy and children’s cardiac function and structure: A systematic review and meta-analysis of evidence from human studies. PLOS ONE. 2022;17(11):e0275236. doi:10.1371/journal.pone.0275236

30. Ullmo S, Vial Y, Di Bernardo S, et al. Pathologic ventricular hypertrophy in the offspring of diabetic mothers: a retrospective study. Eur Heart J. 2007;28(11):1319–1325.

31. Helle EIT, Biegley P, Knowles JW, et al. First Trimester Plasma Glucose Values in Women without Diabetes are Associated with Risk for Congenital Heart Disease in Offspring. J Pediatr. 2018;195:275–278.

32. Suzuki N, Svensson K, Eriksson UJ. High glucose concentration inhibits migration of rat cranial neural crest cells in vitro. Diabetologia. 1996;39(4):401–411. doi:10.1007/s001250050459

33. Basu M, Garg V. Maternal hyperglycemia and fetal cardiac development: Clinical impact and underlying mechanisms. Birth Defects Research. 2018;110(20):1504–1516. doi:10.1002/bdr2.1435

34. Ryckman KK, Spracklen CN, Smith CJ, Robinson JG, Saftlas AF. Maternal lipid levels during pregnancy and gestational diabetes: a systematic review and meta-analysis. BJOG: An International Journal of Obstetrics & Gynaecology. 2015;122(5):643–651. doi:10.1111/1471-0528.13261

35. White SL, Pasupathy D, Sattar N, et al. Metabolic profiling of gestational diabetes in obese women during pregnancy. Diabetologia. 2017;60(10):1903–1912.

36. Mokkala K, Vahlberg T, Pellonperä O, Houttu N, Koivuniemi E, Laitinen K. Distinct Metabolic Profile in Early Pregnancy of Overweight and Obese Women Developing Gestational Diabetes. J Nutr. 2020;150(1):31–37.

37. Huida J, Ojala T, Ilvesvuo J, Surcel HM, Priest JR, Helle E. Maternal first trimester metabolic profile in pregnancies with transposition of the great arteries. Birth Defects Res. Published online December 22, 2022. doi:10.1002/bdr2.2139

38. Cao L, Du Y, Zhang M, et al. High maternal blood lipid levels during early pregnancy are associated with increased risk of congenital heart disease in offspring. Acta Obstet Gynecol Scand. 2021;100(10):1806–1813.

39. Miao Y, Tian L, Martin M, et al. Intrinsic Endocardial Defects Contribute to Hypoplastic Left Heart Syndrome. Cell Stem Cell. Published online August 10, 2020. doi:10.1016/j.stem.2020.07.015

40. Morgan SC, Relaix F, Sandell LL, Loeken MR. Oxidative stress during diabetic pregnancy disrupts cardiac neural crest migration and causes outflow tract defects. Birth Defects Res A Clin Mol Teratol. 2008;82(6):453–463.

41. Grossfeld P, Nie S, Lin L, Wang L, Anderson RH. Hypoplastic Left Heart Syndrome: A New Paradigm for an Old Disease? J Cardiovasc Dev Dis. 2019;6(1). doi:10.3390/jcdd6010010

42. Wahabi HA, Alzeidan RA, Esmaeil SA. Pre-pregnancy care for women with pregestational diabetes mellitus: a systematic review and meta-analysis. BMC Public Health. 2012;12(1). doi:10.1186/1471-2458-12-792

