## Supplements for "Maternal diabetes and overweight as risk factors for congenital heart defects in offspring - A nationwide register study from Finland"

**Online-only supplementary material**

eMethods

eFigure 1: Flow chart of the selection of study participant

eFigure 2: Prevalence of maternal diabetes during the study period

eFigure 3: The prevalence of maternal obesity and congenital heart defects during the study period

eTable 1: Diagnoseis used in excluding subjects with extracardiac anomalies

eTable 2: CHD subgroup categories were classified according to Atlanta ICD-9 classification

eTable 3: The ICD codes and special reimbursement codes used in the definition of mother’s diabetes from the MBR, the Care Register of Hospital Care and the Social Insurance Institution of Finland.

eTable 4: Tthe comparison between individuals with missing and non-missing data and isolated CHD

eTable 5: The prevalence of CHD in the study population.

eTable 6. The distribution of maternal GDM and no DM between maternal BMI in children with isolated CHD (n=10 254)

eTable 7. The distribution of maternal GDM and no DM between maternal BMI in children without isolated CHD (n=610 497)

**eMethods**

**Detailed description of used registers**

The MBR includes data on live births and on stillbirths of fetuses with a birth weight of at least 500 g or with a gestational age of at least 22 weeks. These data are collected using standardized forms and the register includes information on maternal medical history, pregnancy, and delivery and infant’s early medical history. The RCM contains data on congenital chromosomal and structural anomalies that have been detected or suspected in stillborn and live born infants and fetuses. Major congenital anomalies are defined according to EUROCAT classification^1^. The register includes only cases with at least one major anomaly. The RCM uses Atlanta ICD-9 codes for diagnosis and descriptive diagnosis of congenital anomalies. The CRHC contains in- and outpatient visit data, including diagnoses, in hospitals and health centers.

The statistics on special reimbursements for prescription medicines are based on the reimbursement of outpatient pharmaceutical expenses. All permanent residents of Finland are covered under the Finnish National Health Insurance system and are eligible for reimbursements for the cost of medicines prescribed by a doctor or a dentist.  Entitlement for special medication reimbursement is granted based on a clinician's statement, reviewed by a physician at SII against defined criteria. Special reimbursement is designated with a reimbursement code indicating the medication category (e.g. antidiabetic medication) and an ICD-10 code since 2000. Special reimbursement is not granted for gestational diabetes.

**Definition of diabetes**

Diabetes types were defined stepwise starting with T1DM that ruled out T2DM and GDM, which ruled out GDM. T1DM and T2DM were classified by the respective diagnosis codes in the granted special reimbursement for the costs of insulin purchases or in the CRHC (eTable 3). GDM was defined as previously described using the information in the MBR (pathological OGTT, insulin started during pregnancy, and correct diagnosis codes) ^2^.

During the 2006–2016 study period the screening policy of GDM was changed from risk factor based to comprehensive screening in 2008.Current Care Guidelines recommended a 75 g two-hour oral glucose tolerance test at weeks 24-28 for all women except those with very low GDM risk. For women with high risk, an additional oral glucose tolerance test was recommended at 12-16 weeks. The 2008 guideline, which was gradually implemented over the following years, also unified the cut-off concentrations for venous plasma glucose indicating GDM to ≥5.3 mmol/l at baseline (fasting glucose), ≥10.0 mmol/l at 1 hour after glucose intake or ≥8.6 mmol/l at 2 hours after glucose intake ^3^.

**Definition of maternal and paternal education**

Data on the mother’s and father’s highest completed education during the study period were categorised according to the International Standard Classification of Education 2011 (ISCED) as low (ISCED classes 0 to 2), intermediate (ISCED classes 3 to 5), high (ISCED classes 6 to 8) or missing ^4^ .

**References**

1.	Greenlees R, Neville A, Addor MC, et al. Paper 6: EUROCAT member registries: Organization and activities. *Birth Defects Research Part A: Clinical and Molecular Teratology*. 2011;91(S1):S51-S100. doi:[10.1002/bdra.20775](http://dx.doi.org/10.1002/bdra.20775)

2.	Pukkila J, Mustaniemi S, Lingaiah S, et al. Increased Oral Care Needs and Third Molar Symptoms in Women with Gestational Diabetes Mellitus: A Finnish Gestational Diabetes Case-Control Study. *Int J Environ Res Public Health*. 2022;19(17). doi:[10.3390/ijerph191710711](http://dx.doi.org/10.3390/ijerph191710711)

3.	Keikkala E, Mustaniemi S, Koivunen S, et al. Cohort Profile: The Finnish Gestational Diabetes (FinnGeDi) Study. *Int J Epidemiol*. 2020;49(3):762-763g.

4.	UNESCO Institute for Statistics. International Standard Classification of Education ISCED 2011. UNESCO Institute for Statistics 2012.

eTable 1. Diagnoses according to Atlanta ICD-9 codes used in excluding subjects with extracardiac anomalies

| **Subject groups** | **Atlanta ICD-9 diagnosis codes** |
| --- | --- |
| Excluded CHD diagnosis | 745410, 745520, 746870, 746887, 747000, 747230, 747260, 747400, 747410, 747430, 759005, 745500 |
| Excluded chromosomal alterations | 7580, 7581, 7582, 758000, 758020, 758040, 758010, 758030, 758090, 758008, 758098, 758200, 758220, 758240, 758210, 758230, 758290, 758295, 758208, 758298, 758100, 758120, 758110, 758130, 758190, 758108, 758198, 7585, 758510, 758520, 758530, 758540, 758585,758586, 758580, 758500, 758590, 7583, 758360, 758320, 758310,758300, 758330, 758340, 758350, 758380, 758390, 7584, 758400,758600, 758610, 758690, 748850, 758810, 758800, 758860, 7587, 758700, 758710, 758790, 758840, 758820, 758830, 758890, 758900, 758880, 758990 |
| Excluded syndromes / teratogenic syndromes | 742310, 742280, 742480, 742800, 743480, 744880, 759005, 745410, 754010, 753000, 753160, 753180,  755800, 755880, 755810, 756040, 756046, 756110, 756400, 756550, 756410, 756720, 756850, 756800, 757300, 757346, 756045, 757330, 237200, 237700, 759500, 759600, 759610, 759620, 759630, 760700, 760750, 760710, 756030, 756057, 756055, 756060, 756065, 524080, 352600, 759810, 759820, 759800, 759899, 759840, 756830, 759700, 759870, 759860, 759680, 759070, 759340, 279100, 279110, 279910, 758370, 255200, 257800, 742810, 752086, 757520, 759881, 759890, 858610, 59890, 959840, 760718 |
| Extracardiac malformations classified as major anomalies in Registry of Congenital Malformations were excluded to investigate solely isolated congenital heart defects | 216902, 228000, 228010, 228100, 228101, 238000, 238010, 238040, 238080, 243990, 425300, 524000, 658800, 740020, 740080, 741, 741000, 741010, 741030, 741050, 741060, 741086, 741087, 741090, 741920, 741930, 741940, 741980, 741985, 741990, 742000, 742080, 742085, 742086, 742090, 7421, 742100, 742200, 742210, 742220, 742230, 742240, 742250, 742260, 742270, 742290, 742300, 742320, 742380, 742385, 742390, 742400, 742410, 742420, 742485, 742490, 742500, 742520, 742530, 742540, 742580, 742880, 742900, 742910, 742990, 743000, 743010, 743100, 743200, 743210, 743300, 743310, 743320, 743326, 743330, 743340, 743380, 743390, 743400, 743410, 743420, 743430, 743440, 743490, 743500, 743510, 743520, 743530, 743535, 743580, 743590, 743600, 743636, 743640, 743660, 743670, 743680, 743690, 7438, 743800, 743900, 744000, 744010, 744020, 744030, 744090, 744110, 7442, 744210, 744280, 744480, 744500, 744800, 744810, 744881, 746002, 746180, 746310, 746680, 746880, 746882, 747325, 747440, 747450, |
| **Subject groups** | **Atlanta ICD-9 diagnosis codes** |
| Extracardiac malformations classified as major anomalies in Registry of Congenital Malformations were excluded to investigate solely isolated congenital heart defects | 747480, 747490, 7476, 747600, 747610, 747620, 747630, 747640, 747680, 747690, 747800, 747810, 747880, 747900, 748, 748000, 748100, 748120, 748180, 748181, 748208, 748209, 748300, 748310, 748330, 748340, 748350, 748380, 748385, 748390, 748400, 748410, 748480, 748500, 748510, 748520, 748580, 7486, 748620, 748625, 748690, 748810, 748880, 748900, 749, 7490, 749020, 749060, 749070, 749090, 7491, 749100, 749110, 749120, 749170, 749190, 7492, 749200, 749210, 749290, 750110, 750120, 750140, 750180, 7502, 750210, 750230, 750250, 750280, 750300, 750310, 750320, 750330, 750340, 750380, 750420, 750430, 750480, 750580, 750700, 750730, 750750, 750780, 750800, 750900, 750910, 750920, 750990, 751010, 751100, 751110, 751120, 751190, 751200, 751210, 751220, 751230, 751240, 751300, 751310, 751330, 751340, 751400, 751410, 751420, 751490, 751495, 751500, 751520, 751530, 751540, 751550, 751560, 751580, 751590, 751600, 751610, 751620, 751630, 751640, 751650, 751660, 751670, 751700, 751710, 751720, 751730, 751780, 751820, 751880, 7519, 751900, 752000, 752010, 752080, 752085, 752088, 752190, 752200, 752300, 752310, 752380, 752390, 752400, 752410, 752420, 752480, 752490, 752530, 752600, 752605, 752606, 752607, 752610, 752620, 752621, 752625, 752626, 752627, 752700, 752710, 752730, 752790, 7528, 752800, 752820, 752830, 752840, 752850, 752860, 752862, 752865, 752880, 752900, 753001, 753009, 753010, 753100, 753110, 753120, 753130, 753140, 753150, 7532, 753200, 753210, 753220, 753290, 753300, 753310, 753320, 753330, 753380, 753400, 753410, 753420, 753480, 753485, 753490, 753500, 753501, 7536, 753600, 753610, 753620, 753630, 753680, 753690, 7537, 753700, 753710, 753790, 753800, 753820, 753830, 753840, 753850, 753860, 753880, 753900, 753920, 753990, 754001, 754030, 754050, 754061, 754080, 7541, 754100, 754280, 7543, 754300, 754310, 754500, 754580, 754735, 754780, 7548, 754830, 754880, 7550, 755005, 755006, 755007, 755010, 755020,755030, 755090, 755095, 755096, 7551, 755100, 755110, 755120, 755130, 755131, 755190, 755191, 755192, 755193, 755194, 755195, 755196, 755199, 755200, 755210, 755220, 755230, 755240, 755250, 755260, 755270, 755280, 755290, 755300, 755330, 755340, 755350, 755365, 755366, 755380, 755390, 755410, 755480, 755500, 755510, 755520, 755525, 755526, 755530, 755536,755540, 755550,755555, 755556, 755580, 755585, 755610, 755620, 755630, 755631, 755640, 755647, 755650, 755660, 755665, 755666, 755667, 755670, 755680, |
| **Subject groups** | **Atlanta ICD-9 diagnosis codes** |
| Extracardiac malformations classified as major anomalies in Registry of Congenital Malformations were excluded to investigate solely isolated congenital heart defects | 755685, 755881, 755900, 756005, 756006, 756010, 756020, 756050, 756080, 756081, 756085, 756090, 7561, 756120, 756130, 756140 , 756145, 756150, 756155, 756156, 756160, 756165, 756166, 756170, 756179, 756180, 756185, 756190, 756300, 756310, 756320, 756330, 756340, 756350, 756380, 756390, 756420, 756430, 756440, 756445, 756446, 756447, 756451, 756460, 756470, 756480, 756490, 756500, 756540, 756560, 756575, 756600, 756610, 756615, 756616, 756617, 756620, 756680, 756700, 756710, 756780, 756790, 756810, 756860, 756880, 756900, 756920, 756990, 757, 757000, 757110, 757115, 757190, 757195, 757196, 757280, 7573, 757320, 757340, 757345, 757350, 757360, 757380, 757382, 757390, 757391, 757395, 757400, 757480, 757500, 7576, 757600, 757620, 757630, 757800, 757990, 758850, 759, 759000, 759010, 759040, 759050, 759080, 759100, 759110, 759180, 759210, 759220, 759240, 759290, 7593, 759300, 759320, 759330, 759390, 7595, 7596, 759690, 7597, 7598, 759900, 759991, 771100, 771140, 771210, 771220, 778000 |

eTable S2. CHD subgroups classified according to Atlanta ICD-9 codes

| **Congenital heart defects subgroup** | **Atlanta ICD-9 codes** |
| --- | --- |
| Complex | 746, 7453, 7457, 745180,745190, 745300, 745420, 745610, 745700, 746800, 746810, 746820, 746887, 746888, 746889 |
| TGA, transposition of great arteries | 7451, 745100, 745110, 745120 |
| LVOTO, left ventricular outflow tract obstruction | 7463, 7464, 7365, 7466, 7467, 7471, 746300, 746400, 746401, 746480, 746481, 746490, 746500, 746505, 746580, 746581, 756600, 746601, 746700, 756900, 746901, 747100, 747110, 747190, 747200, 747210, 747220 |
| RVOTO, right ventricular outflow tract obstruction | 7452, 7460, 7461, 7462, 7473, 745200, 745210, 746000, 746010, 746020, 746021, 746080, 746081, 746090, 746100, 746105, 746106, 746181, 746200, 746830, 747130, 747200, 747300, 747310, 747320, 747380, 747390 |
| Pulmonary venous anomalies | 747420, 747430 |
| Septal - others | 745, 7450, 7456, 7456, 7458, 7459, 745000, 745010, 745400, 745620, 745630, 745680, 745690, 745800, 745900 |
| VSD - ventricular septal defect | 7454, 74548, 745480, 745481, 745482, 745483, 745484, 745485, 745486, 745487, 745488, 745490, 745491, 745492, 745493, 745494, 745495, 745496, 745498 |
| ASD - atrial septal defect | 745600, 7455, 745590, 745510, 745580 |
| Thoracic arteries and veins | 7472, 746885, 747200, 747215, 747240, 747250, 747270, 747280, 747290, 747330, 747340, 747380, 748685 |

eTable 3. The ICD codes used in the definition of mother’s diabetes^a^

| **Definition** | **ICD codes** |
| --- | --- |
| T1DM | ICD-9: 250*B  ICD-10: E10, O24.0 |
| T2DM / other type of diabetes | ICD-9: 250*A, 250*C, 250*X  ICD-10: E11, E12, E13, E14, O24.1, O24.2, O24.3 |
| GDM | ICD-9: 6480A, 6488A  ICD-10: O24.4, O 24.9 |

Abbreviations: T1DM, type 1 diabetes; T2DM, type 2 diabetes or other diabetes; GDM, gestational diabetes.

^A^ The ICD codes were used from MBR, the Care Register of Hospital Care and the Register of Special Reimbursement (special reimbursement for insulin).

|  | **No. (% of whole study population)** |
| --- | --- |
| Isolated CHD | 10 254 (1.65) |
| Complex | 47 (0.01) |
| TGA | 133 (0.02) |
| LVOTO | 1105 (0.18) |
| RVOTO | 1101 (0.18) |
| Pulmonary venous anomalies | 36 (0.01) |
| Septal - others | 411 (0.07) |
| VSD | 6468 (1.04) |
| ASD | 803 (0.13) |
| Thoracic veins and arteries | 128 (0.02) |

eTable 4. The prevalence of isolated CHDs^a^ in the study population (n=620 751)

Abbreviations: CHD, congenital heart defect; TGA, transposition of great arteries; LVOTO, left ventricle outflow tract obstruction; RVOTO, right ventricle outflow tract obstruction; VSD, ventricular septal defect; ASD, atrial septal defect.

^a^See specified list of diagnose codes in eTable 1.

eTable 5. The presence of maternal GDM and no DM according to BMI in mothers of children with and without isolated CHD^a^

| **Mothers of children with isolated CHD** | **No DM (n = 8469)** | **GDM (n = 1524)** |
| --- | --- | --- |
| BMI <18.5 kg/m^2^, No. (%) | 375 (95.4) | 10 (2.5) |
| BMI 18.5-24.9 kg/m^2^, No. (%) | 5555 (89.7) | 505 (8.2) |
| BMI 25.0-29.9 kg/m^2^, No. (%) | 1655 (76.0) | 459 (21.1) |
| BMI ≥30 kg/m^2^, No. (%) | 711 (54.8) | 534 (41.1) |
| **Mothers of children without isolated CHD** | **No DM (n = 520 460)** | **GDM (n = 85 503)** |
| BMI <18.5 kg/m^2^, No. (%) | 20815 (95.0) | 1009 (4.6) |
| BMI 18.5-24.9 kg/m^2^, No. (%) | 342 982 (92.1) | 27 527 (7.4) |
| BMI 25.0-29.9 kg/m^2^, No. (%) | 101 607 (78.2) | 27 256 (21.0) |
| BMI ≥30 kg/m^2^, No. (%) | 44 247 (60.0) | 28 326 (38.4) |

Abbreviations: CHD, congenital heart defect, GDM, gestational diabetes; DM diabetes; BMI, body mass index.

^a^Data of mothers BMI status were missing in 12 454 (2.0%) cases.

eTable 6. The comparison of isolated CHD between participants with missing data and non-missing data^a^

| Characteristics | Isolated CHDs (n=10 254) | Healthy controls (n=610 497) | p-value |
| --- | --- | --- | --- |
| Missing data of mother’s smoking, No. (%) | 258 (1.7) | 14 587 (98.3) | .40 |
| Non-missing data of mother’s smoking, No. (%) | 9996 (1.7) | 595 910 (98.4) |  |
| Missing data of mother’s BMI status, No. (%) | 194 (1.6) | 12 260 (98.4) | .43 |
| Non-missing data of mother’s BMI status, No. (%) | 10 060 (1.6) | 598 237 (98.4) |  |
| Missing data of mother’s education level, No. (%) | 1060 (1.7) | 61 498 (98.4) | .38 |
| Non-missing data of mother’s education level, No. (%) | 9194 (1.7) | 548 999 (98.3) |  |
| Missing data of father’s education level, No. (%) | 1575 (1.7) | 96 791 (98.4) | .18 |
| Non-missing data of father’s education level, No. (%) | 8679 (1.7) | 513 706 (98.3) |  |

Abbreviations: No, number.

^a^The statistical significance was reached with p-value <.05.

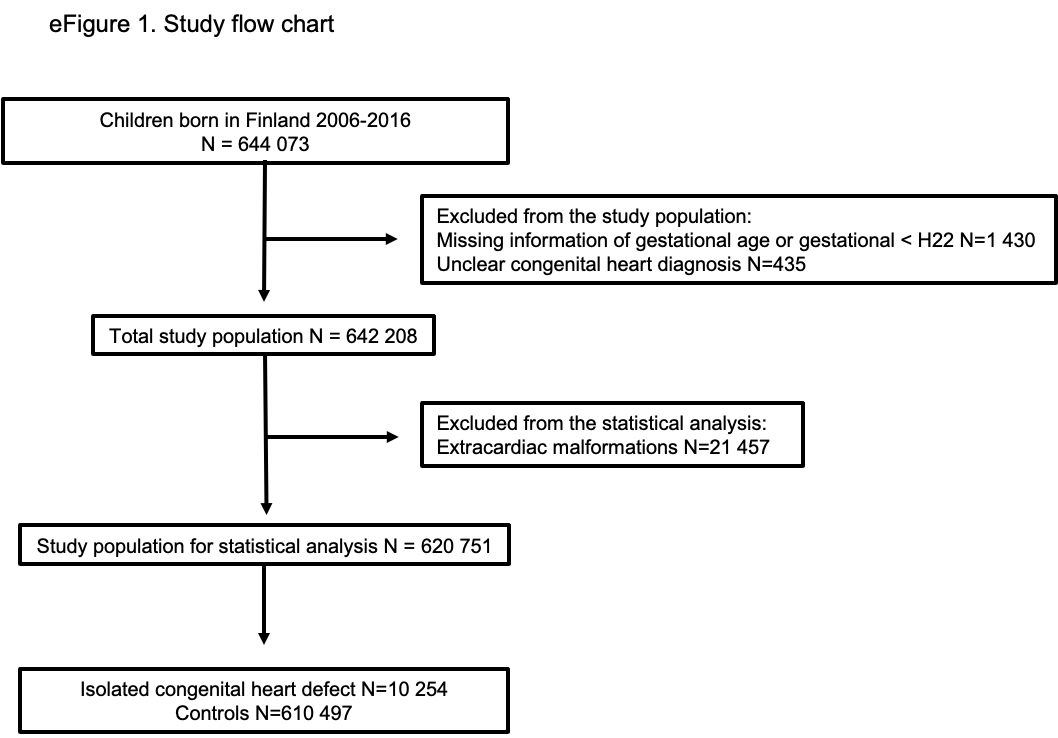

eFigure 2. The prevalence of maternal diabetes and congenital heart defects during the study period
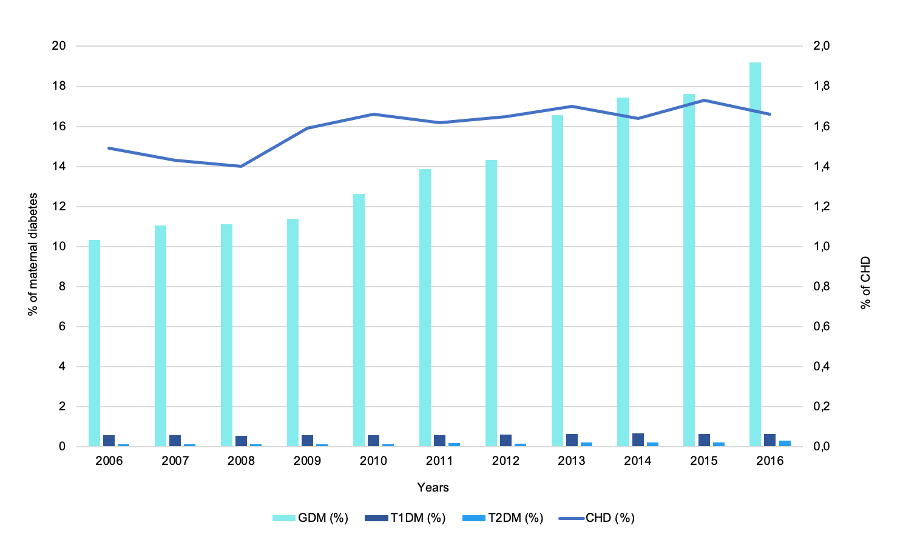

Abbreviations: GDM, gestational diabetes mellitus; T1DM, type 1 diabetes; T2DM, type 2 diabetes or other diabetes; CHD, congenital heart defect.

eFigure 3. The prevalence of maternal obesity and congenital heart defects during the study period
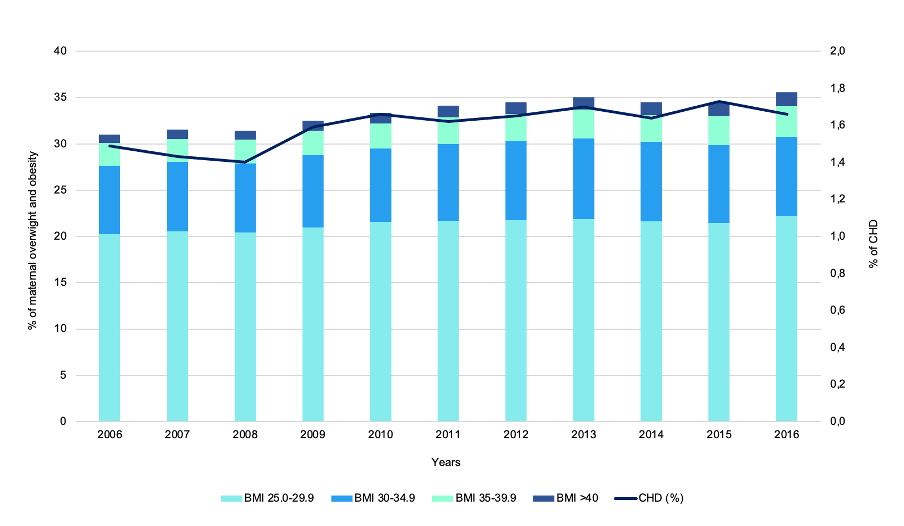

Abbreviations: BMI, body mass index; CHD, congenital heart defect.
